# Perinatal colonization with extended-spectrum beta-lactamase-producing and carbapenem-resistant Gram-negative bacteria among home births in Bangladesh

**DOI:** 10.1101/2025.05.14.25327637

**Authors:** Gregory S. Wu, Hafsa Hossain, Mohammed Badrul Amin, Shahana Parveen, Mohammad Aminul Islam, Ajrin Sultana Sraboni, Abu Faisal Md. Pervez, Dilruba Zeba, Emily S. Gurley, Stephen Luby, Ashley Styczynski

## Abstract

**Background:** Hospital-acquired neonatal infections are increasingly caused by antibiotic-resistant bacteria. It is unknown to what extent home-based births, which account for nearly a third of deliveries in Bangladesh, may also result in exposure to antibiotic-resistant bacteria.

**Methods:** We enrolled mothers who delivered at home and their newborns from a rural community in Bangladesh during April-June, 2022. Within 2-7 days after delivery, we collected vaginal and rectal swabs from mothers and rectal swabs from the newborns. Swabs were cultured on chromogenic culture media selective for extended-spectrum β-lactamase-producing bacteria (ESBL-PB) and carbapenem-resistant bacteria (CRB). Demographic and risk factor data were collected via surveys. Birth attendants who facilitated the deliveries were interviewed regarding infection prevention practices. We performed descriptive analyses to identify potential risk factors associated with colonization.

**Results:** Of the 50 mothers enrolled, the median age was 23 years (range 18 to 26). Thirty-eight (76%) mothers had at least one antenatal care visit. Only one mother reported hospitalization during pregnancy, and 4 reported antibiotic use during pregnancy. Following delivery, 47 (94%) mothers were colonized with ESBL-PB, and 37 (74%) were colonized with CRB. Of the newborns, 36 (72%) were colonized with ESBL-PB, and 27 (54%) were colonized with CRB. No associations were found with any perinatal exposures, though all households reported incomes below the international poverty level. Of the 9 birth attendants were able to be interviewed, 7 (78%) reported performing hand hygiene before delivery, and 8 (89%) reported glove use during the delivery. Attendants reported cleaning equipment shared across deliveries with soap and water and using boiled water for delivery (89%, n=8).

**Conclusions:** Women and newborns in this rural population were frequently colonized with both ESBL-PB and CRB following home deliveries. This demonstrates the importance of community-based antibiotic-resistant bacterial transmission and need for further understanding community exposures driving antibiotic resistance.

## Background

Antimicrobial resistance (AMR) is a critical public health threat that disproportionately affects low- and middle-income countries [1]. The threat is particularly profound for vulnerable populations such as newborns, whose immature immune systems and microbiomes are less able to effectively protect them from invasive infections. Neonatal sepsis is increasingly driven by antibiotic resistant bacteria, which has hindered progress in reaching the United Nations sustainable development goal 3 of reducing newborn mortality [2]. Moreover, poverty and infections caused by antibiotic-resistant bacteria may interact to further increase the risk for adverse infectious outcomes, as neonatal sepsis has also been linked with lower household wealth metrics [3].

Colonization with a bacterial pathogen typically precedes infection and puts individuals at higher risk of developing an infection caused by the same bacteria [4–6]. This relationship between bacterial colonization and infection has been well documented in the hospital setting but has also been demonstrated in the community [6,7]. In the case of Group B streptococcus (GBS), maternal colonization has been associated with 9-fold increased risk for neonatal infections [8]. This has resulted in implementation of screening and antibiotic prophylaxis for pregnant women who are colonized with GBS in many high-resource settings [9–11]. However, in low-resource settings, Gram negative bacteria, particularly antibiotic-resistant Gram negative bacteria, are increasingly a dominant cause of neonatal infections compared to GBS [12–14]. Although less is known about the relationship between maternal colonization with Gram negative bacteria and neonatal outcomes, recent studies identified an association between maternal colonization with antibiotic-resistant Gram negative bacteria and a variety of adverse outcomes, including premature birth, preterm premature rupture of membranes, perinatal asphyxia, and breech birth [8,15].

Compared to adults, newborns lack well-developed, diverse microbiomes, which can provide resistance to colonization by pathogens and infection. As such, colonization at birth largely reflects the environment they are born into. Traditionally, the newborn microbiome is thought to be heavily shaped by the maternal gut and vaginal microbiome, particularly among vaginally-delivered infants [16–18]. This pattern can be disrupted among newborns undergoing Cesarean deliveries (C-section), exposed to antibiotics (either directly or indirectly through maternal administration pre-delivery), or experiencing prolonged hospitalization [19–21]. These nuances in care and the birth environment can play a crucial role in shaping the newborn’s gut microbiome.

A key strategy for reducing maternal and infant mortality has been shifting from home-based deliveries to facility-based deliveries, ensuring access to lifesaving intrapartum services as well as antenatal and postnatal care [22]. As a result, substantial improvements have been made: since 1990, maternal and neonatal mortality have been cut in half [23,24]. Although these improvements may not be solely attributable to facility-based deliveries, current strategies continue to emphasize the important role that healthcare plays in improving birth outcomes [25].

However, it is unknown to what extent this shift toward facility-based deliveries may be influencing the exposure of newborns to antibiotic-resistant bacteria. In a previous report, we found a high prevalence of colonization with extended-spectrum beta-lactamase-producing bacteria (ESBL-PB) and carbapenem-resistant bacteria (CRB) among newborns and pregnant mothers undergoing delivery in a hospital setting in Bangladesh [26]. Maternal colonization with ESBL-PB increased from 77% on admission to 98% following delivery, while colonization with CRB increased from 15% to 89%. Among the newborns, 89% were found to be colonized with ESBL-PB and 72% with CRB. Paired with high rates of antibiotic usage and C-sections as well as frequent contamination of hospital environments with these pathogens, these findings implicated the hospital setting in the transmission of antibiotic-resistant bacteria to newborns. Similarly, a large multicenter study of facility-based deliveries found very few genetically related antibiotic-resistant bacteria between mother and newborn pairs, indicating horizontal transmission as a key exposure pathway [15].

Although hospital-based deliveries have been increasing in Bangladesh, one-third of deliveries are still taking place in the home [27]. Roughly half of these deliveries are attended by a medically trained provider, which includes nurses, midwives, paramedics, family welfare visitors, community skilled birth assistants, and sub-assistant community medical officers. In contrast with facility-based deliveries, little is known about the infection control practices of home-based deliveries. Additionally, the exposure to antibiotic-resistant bacteria may vary by birth setting. This is supported by findings of increased newborn colonization with CRB among those born in a study facility compared with those born outside a study facility, which included home births [15]. This study was conducted to evaluate ESBL-PB and CRB colonization patterns among newborns delivered at home and assess perinatal exposures and infection control practices that may contribute to colonization with antibiotic-resistant bacteria in home births.

## Methods

### Participant enrollment

For this study, we enrolled mother and newborn pairs between April 4, 2022 and June 28, 2022 who had undergone home-based deliveries from a rural community in Baliakandi, a sub-district under Rajbari district, Bangladesh. Mothers were identified through an ongoing birth tracking notification system as part of the Child Health and Mortality Prevention Surveillance (CHAMPS) program. Following the notification of a birth, we contacted families via phone to explain the study’s objectives. After initial agreement to participate, we visited the household between 2 and 7 days after delivery. We obtained written informed consent from all adult study participants. Consent for newborn enrollment was provided by the parents and/or legal guardians. A trained female study physician with the help of a female field assistant collected vaginal and rectal swabs from the mothers and rectal swabs from the newborns. We also conducted interviews with mothers to obtain additional information on demographics, prenatal history, and perinatal exposures.

### Bacterial culturing of swab samples

Both rectal and vaginal swabs were placed in Amies media (BactiSwab™, ThermoScientific, Waltham, MA, USA) and maintained at 4-8°C and transported to the icddr,b lab in Dhaka for processing within 24 hours. Swabs were inoculated onto CHROMagar™ ESBL and CHROMagar mSuperCARBA (CHROMagar, Paris, France). Colonization was determined by the presence of bacterial growth on either of the culture plates after overnight incubation at 37°C. Mothers were considered to be colonized with ESBL-PB or CRB if there was growth from either the vaginal or rectal swab on the respective media; newborns were considered to be colonized with ESBL-PB or CRB if there was growth from the rectal swab on the respective media.

### Birth attendant interviews

To determine home birthing practices, we conducted interviews with birth attendants who attended deliveries of the enrolled mother-newborn pairs. We collected an initial list of 12 attendants from enrolled mothers who conducted their deliveries. Attendants were selected based on our research team’s ability to locate them and their willingness to participate in interviews. A total of nine birth attendants were interviewed. A semi-structured questionnaire was used to collect data on their prior training, delivery practices, infection prevention procedures, and interactions with healthcare facilities.

### Statistical analysis

We performed descriptive analysis to characterize participant demographics, colonization prevalence, and birth attendant practices. To identify potential risk factors for colonization, we performed univariate analysis. We used Fisher’s exact test to calculate p-values, 95% confidence intervals, and odds ratios for each variable. Statistical significance was determined by a p-value less than 0.05. Categories where all participants responded identically were excluded as the Fisher’s exact test could not be performed. For variables in which one of the categories was zero, we applied the Haldane-Anscombe correction to calculate the odds ratio [28,29].

## Results

### Demographics of enrolled participants

We enrolled 50 mothers and newborns. All deliveries were non-assisted (i.e., without use of instruments such as forceps or vacuum cups), live vaginal births that took place in the household. The median age of the mothers was 23 years (range 18-26) (Table 1). The median monthly household income was $118 USD, and the median household size was 6 people. This amounts to $0.66 USD per person per day, which meant that all households fell below the World Health Organization poverty level ($1.90 USD per person per day at the time of the study). Approximately half (46%, n=23) of mothers reported their highest education level as primary school or lower, and 48 (96%) identified homemaker as their occupation. All participants reported using a pit latrine, either shared or private, as their toilet type, and all reported handwashing facilities with soap and water at their residence. Additionally, all mothers reported physical contact with domestic animals during their pregnancy.

**Table 1:** Demographics of mothers undergoing home-based deliveries, Baliakandi, Bangladesh, 2022 (N=50)

| Community Variable | n | % or range |
| --- | --- | --- |
| Median age of women (years) | 23 | 18-26 |
| Highest education completed |  |  |
| No formal education | 9 | 18 |
| Primary school | 14 | 28 |
| Secondary school | 20 | 40 |
| College (equivalent to the final 2 years of high school) | 6 | 12 |
| Bachelor's degree | 1 | 2 |
| Occupation |  |  |
| Homemaker | 48 | 96 |
| Tailor | 1 | 2 |
| Salaried government job | 1 | 2 |
| Domestic animal contact during pregnancy |  |  |
| Animal residence inside the home | 35 | 70 |
| Animal residence outside the home | 50 | 100 |
| Monthly income |  |  |
| Median monthly income (USD) | 118 | 47-212 |
| Median household size (persons) | 6 | 3-8+ |
| Main water source - tube well | 49 | 98 |
| Water treatment - no treatment | 49 | 98 |
| Toilet type |  |  |
| Shared pit latrine | 24 | 48 |
| Private pit latrine | 26 | 52 |
| Handwashing facilities – yes | 50 | 100 |
| Nulliparous at enrollment | 18 | 36 |
| Antenatal care visits |  |  |
| None | 12 | 24 |
| 1-3 visits | 30 | 60 |
| 4 or more visits | 8 | 16 |
| Pregnancy complications, self-reported |  |  |
| No complication | 34 | 68 |
| High blood pressure | 1 | 2 |
| Underweight | 1 | 2 |
| Anemia | 12 | 24 |
| Urinary infection | 6 | 12 |
| Timing of delivery |  |  |
| Preterm (<37 weeks) | 3 | 6 |
| Full term (37-42 weeks) | 46 | 92 |
| Post term (>42 weeks) | 1 | 2 |

One-third of the women had no prior pregnancies. Thirty-eight (76%) mothers reported at least some antenatal care, but 42 (84%) mothers had less than 4 antenatal care visits prior to delivery. Only 4 (8%) mothers reported using antibiotics in the 30 days prior to delivery. The most commonly self-reported pregnancy complications were anemia (n=12, 24%) and urinary tract infection (n=6, 12%). Most deliveries occurred at term (n=46, 92%). One participant was hospitalized during her pregnancy, but none of the enrolled mothers or babies were hospitalized after delivery.

### Colonization prevalence and associated risk factors

ESBL-PB colonization was more frequent among mothers compared with newborns (Figure 1). Forty-seven (94%) mothers were colonized with ESBL-PB, while 37 (74%) were colonized with CRB. Thirty-seven (74%) mothers were colonized with both. Of the newborns, 36 (72%) were colonized with ESBL-PB, and 27 (54%) were colonized with CRB. Twenty-six (48%) newborns were colonized with both.

**Figure 1:**
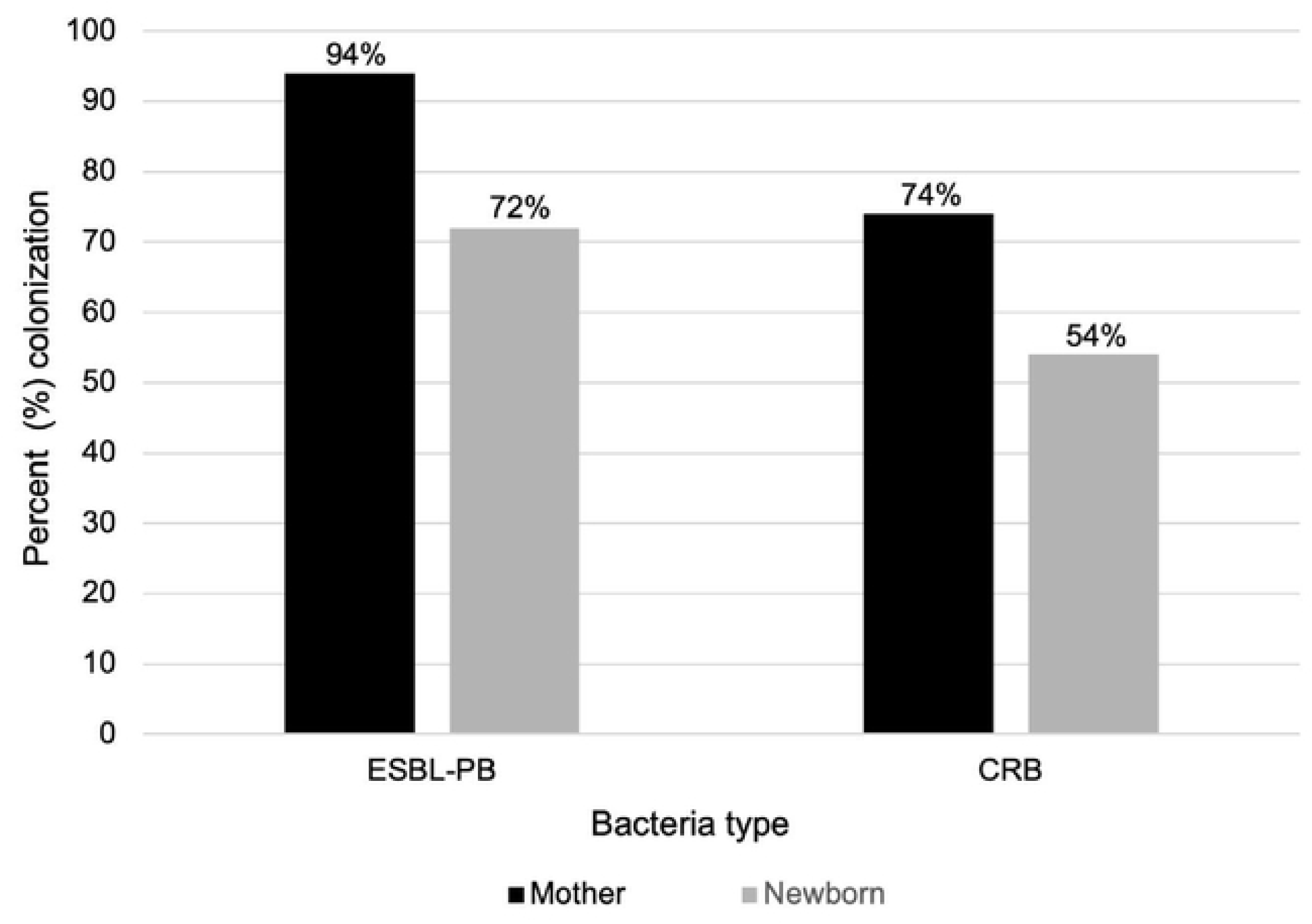
Colonization prevalence among mothers and newborns following home-based delivery, Baliakandi, Bangladesh, 2022 (N=50) Colonization was determined based on bacterial growth on selective media from rectal and/or vaginal swabs. ESBL-PB = extended-spectrum beta-lactamase-producing bacteria; CRB = carbapenem-resistant bacteria

Mothers with ESBL-PB colonization compared to those without ESBL-PB colonization were more likely to report a household income at or below the median (100% vs 75%) and 5 or fewer people living in the household (100% vs 89%) (Table 2). They were also more likely to have contact with domestic animals inside the home (97% vs 87%), a shared vs private pit latrine (100% vs 88%), and no medication use in the prior 30 days (100% vs 88%). However, none of these differences were statistically significant. The most common animals contacted inside the home were chickens and cats (Supplementary Table 1).

**Table 2:** Comparison of maternal colonization patterns following home based delivery across community variables, Baliakandi, Bangladesh (N=50)

|  | ESBL-PB colonization |  |  |  | CRB colonization |  |  |  |
| --- | --- | --- | --- | --- | --- | --- | --- | --- |
| Community Variable | <u>Yes</u><br>N (%) | <u>No</u><br>N (%) | OR (95% CI) | p<br>value | <u>Yes</u><br>N (%) | <u>No</u><br>N (%) | OR (95% CI) | p<br>value |
| Highest education completed |  |  |  |  |  |  |  |  |
| Primary and below | 22 (96%) | 1 (4.3%) | Ref | - | 15 (65%) | 8 (35%) | Ref | - |
| Secondary and above | 25 (93%) | 2 (7.4%) | 0.57 (0.01-12) | 1.0 | 22 (81%) | 5 (19%) | 2.3 (0.54-11) | 0.22 |
| Household Income |  |  |  |  |  |  |  |  |
| At or below median | 18 (100%) | 0 (0%) | Ref | - | 13 (72%) | 5 (28%) | Ref | - |
| Above median | 24 (75%) | 8 (25%) | 0.08 (0.01-1.4) | 0.54 | 24 (75%) | 8 (25%) | 1.2 (0.24-5.0) | 1.0 |
| Household size |  |  |  |  |  |  |  |  |
| 5 or below | 22 (100%) | 0 (0%) | Ref | - | 19 (86%) | 3 (14%) | Ref | - |
| 6 or more | 25 (89%) | 3 (11%) | 0.16 (0.01-3.3) | 0.25 | 18 (64%) | 10 (36%) | 0.29 (0.044-1.4) | 0.11 |
| Toilet type |  |  |  |  |  |  |  |  |
| Shared pit latrine | 24 (100%) | 0 (0%) | Ref | - | 16 (67%) | 8 (33%) | Ref | - |
| Private pit latrine | 23 (88%) | 3 (12%) | 7.3 (0.40-150) | 0.24 | 21 (81%) | 5 (19%) | 2.1 (0.49-9.7) | 0.34 |
| Water storage |  |  |  |  |  |  |  |  |
| Unsafe water storage | 25 (96%) | 1 (4%) | Ref | - | 18 (69%) | 8 (31%) | Ref | - |
| Safe water storage (closed container, narrow opening) | 22 (92%) | 2 (8%) | 0.45 (0.01-9.1) | 0.60 | 19 (79%) | 5 (21%) | 1.7 (0.39-7.8) | 0.53 |
| Domestic animal contact during pregnancy (inside) |  |  |  |  |  |  |  |  |
| No domestic animal contact | 13 (87%) | 2 (13%) | Ref | 0.21 | 10 (67%) | 5 (33%) | Ref | - |
| Domestic animal contact | 34 (97%) | 1 (3%) | 5.0 (0.24-320) | - | 27 (77%) | 8 (23%) | 1.7 (0.34-7.6) | 0.49 |
| Antenatal care visits |  |  |  |  |  |  |  |  |
| None | 12 (100%) | 0 (0%) | Ref | - | 7 (58%) | 5 (42%) | Ref | - |
| Any | 35 (92%) | 3 (7.9%) | 0.41 (0.02-8.4) | 1.0 | 30 (79%) | 8 (21%) | 2.6 (0.51-13) | 0.26 |
| Pregnancy complications (self-reported) |  |  |  |  |  |  |  |  |
| No complication | 33 (97%) | 1 (2.9%) | Ref | - | 26 (76%) | 8 (24%) | Ref | - |
| Anemia | 10 (83%) | 2 (17%) | 0.14 (0.00-3.0) | 0.14 | 7 (58%) | 5 (42%) | 0.38 (0.077-2.0) | 0.26 |
| Urinary Infection | 6 (100%) | 0 (0%) | 0.58 (0.02-15) | 1.0 | 5 (83%) | 1 (17%) | 1.9 (0.18-96) | 1.0 |
| Medication use within 30 days prior to delivery |  |  |  |  |  |  |  |  |
| No medication use | 26 (100%) | 0 (0%) | Ref | - | 18 (69%) | 8 (31%) | Ref | - |
| Medication use | 21 (88%) | 3 (13%) | 0.12 (0.01-2.4) | 0.10 | 19 (79%) | 5 (21%) | 1.7 (0.39-7.8) | 0.53 |
| Timing of delivery |  |  |  |  |  |  |  |  |
| Full term (37-42 weeks) | 43 (93%) | 3 (6.5%) | Ref | - | 34 (74%) | 12 (26%) | Ref | - |
| Preterm (<37 weeks) | 3 (100%) | 0 (0%) | 0.56 (0.02-13) | 1.0 | 2 (67%) | 1 (33%) | 0.71 (0.06-8.5) | 1.0 |
| Post term (>42 weeks) | 1 (100%) | 0 (0%) | 0.24 (0.01-7.1) | 1.0 | 1 (100%) | 0 (0%) | 1.1 (0.04-28) | 1.0 |

Mothers with CRB colonization compared to those without CRB colonization were more likely to have at least a secondary education (81% vs 65%) and have 5 or fewer people living in the household (86% vs 64%). They also reported more frequently having a private compared with a shared pit latrine (81% vs 67%), having contact with domestic animals inside the home (77% vs 67%), and having had any antenatal care (79% vs 58%), though these differences were also not significant (Table 2).

### Description of home birth practices

Twelve birth attendants were identified by the study participants. Nine birth attendants were able to be contacted and completed interviews, of which 5 were carried out via phone while the other 4 were conducted in person. All birth attendants were female, with a median age of 52 (Supplementary Table 3). Seven (78%) of the birth attendants had completed secondary school. Four (44%) of the birth attendants had received formal midwifery training, 3 (33%) had received informal training, and 2 (22%) had received no training. Three (33%) of the birth attendants reported working under a village doctor or physician, and 2 (22%) reported also attending deliveries in facilities as support staff. The birth attendants averaged 10 deliveries per month (range 0-22) and had an average of 16 years (range 6-35) of experience working as birth attendants.

In preparation for delivery, birth attendants reported frequently laying down blankets or sheets. Four (44%) respondents reported that they disinfect the delivery area. Seven (78%) birth attendants reported performing hand hygiene with soap and water or alcohol-based hand rub before delivery, and 8 (89%) reported glove use during the delivery. Hand hygiene was often reported as being performed before cervical checks, before procedures, and before donning gloves but was infrequently reported as being performed after any of those moments. Respondents frequently reported changing gloves before and after delivering the baby (78%). Exchanging gloves before/after cervical checks or other procedures was inconsistently reported (22%).

Equipment commonly shared across deliveries included scissors, cloth or plastic sheets, blood pressure cuffs, and thermometers. These equipment were typically cleaned with soap and water between deliveries with the exception of blood pressure cuffs that were not cleaned. Most reported that water used during the delivery was boiled (89%). Few of the birth attendants reported performing invasive procedures (i.e., membrane stripping, mechanical cervical ripening, artificial rupture of membranes, or episiotomies). The respondents typically reported cutting the umbilical cord with a sterile scalpel or sterile scissors (78%). After delivery, the respondents reported cleaning the baby’s mouth with gauze (89%) or a finger sweep without gloves (22%). Other immediate newborn care practices included wiping/drying the baby (78%), placing the baby on the mother’s breast (89%), and applying chlorhexidine to the umbilical cord (78%).

## Discussion

Mothers and newborns who underwent home deliveries in a rural setting in Bangladesh were found to have an alarmingly high incidence of ESBL-PB and CRB colonization following delivery. This occurred despite limited interaction with healthcare settings - only one mother reported hospitalization during pregnancy - and rare antibiotic use during pregnancy. This contrasts with studies from high-resource settings that frequently link antibiotic resistance with healthcare settings and antibiotic use [30,31]. The high prevalence of colonization with antibiotic-resistant bacteria in a community setting in the absence of recognized risk factors highlights that the epidemiology of antibiotic-resistant bacteria is context-dependent and varies between high- and low-resource settings.

Colonization with ESBL-PB reported in this study is similar to other reports from Bangladesh, though CRB colonization was found to be higher than in other studies. One study evaluating colonization among children of less than 1-year of age in rural Bangladesh found 82% were colonized with *Escherichia coli* resistant to third generation cephalosporins, and 74% of infants were colonized with ESBL-producing *E. coli* [32]. This study did not report prevalence of CRB. Another study evaluating colonization prevalence in urban Bangladesh reported 78% of adults were colonized with Enterobacterales resistant to third generation cephalosporins but only 9% were colonized with CRB [33]. These estimates align with other reports from South Asia but differ greatly from reports in high-resource settings, where the prevalences of ESBL-PB and CRB colonization are much lower among community populations [34–38]. The difference in CRB colonization is particularly striking given that carbapenem antibiotics are infrequently used in Bangladesh. This could indicate that non-specific mechanisms of antibiotic resistance (e.g., efflux pumps, ampC overproduction) are mediating carbapenem resistance. Alternatively, antibiotic-resistance genes against carbapenems (i.e., carbapenemases) may be co-located on plasmids with other antibiotic-resistance genes where they are being selected for following exposure to non-carbapenem antibiotics or other biocides [39]. Additional genomic analyses could provide further insight into the molecular epidemiology of CRB.

Although colonization of mothers was not evaluated pre-delivery, colonization post-delivery might be expected to be similar to pre-delivery in the absence of any healthcare exposures or interventions. Yet, a previous study conducted among mothers presenting to a hospital in an adjacent community for delivery were found to have substantially lower rates of colonization with antibiotic-resistant bacteria pre-delivery [26]. While the participants in this study were largely similar to the mothers in the hospital cohort, the mothers who underwent home births were of lower income. In addition, all participants in the current study listed pit latrines as their toilet type compared with 79% of mothers in the hospital-based study; the remainder utilized pour-flush toilets, which are generally regarded as more sanitary and hygienic. Moreover, 98% of mothers who underwent home births indicated not treating their water, while only 72% of mothers did in the hospital cohort. These statistics outline a population that is lower income and with less access to water, sanitation, and hygiene (WASH) infrastructure, which may be mediating exposure to antibiotic-resistant bacteria [40].

Although risk factor analyses did not reveal any clear perinatal exposures linked with colonization, all participants met criteria for living in poverty, which has been directly associated with increased risk for colonization or infection with antibiotic-resistant bacteria [41]. A prior study evaluating risk factors for colonization among pregnant women in India identified that low socioeconomic status was associated with increased risk for urinary colonization with ESBL-PB [42]. Similarly, more global analyses have linked increased antibiotic resistance with lower development indices, such as governance, health expenditure, and infrastructure [43,44]. Additionally, WASH is a poverty-related factor that has been linked with increased antibiotic resistance [40,43,45]. In this study, participants reported primarily relying on pit latrines and tubewells. While these are frequently classified as improved WASH infrastructure, improved infrastructure does not always translate to a lack of exposure to fecal pathogens [46]. Additionally, because humans are a major reservoir for antibiotic-resistant bacteria, increased fecal exposure likely parallels increased exposure to antibiotic-resistant bacteria [47]. Several studies have found fecal bacterial contamination in tubewells, regardless of depth, which has been attributed to potential breaks in the structures [48,49]. A meta-analysis of studies from LMICs found fecal contamination to be higher from boreholes compared with piped water (43% vs. 25%) [50]. Similarly, another systematic review found 3.45-times higher odds of fecal contamination among self-supplied water (primarily comprised of boreholes) compared with piped public water in LMICs [51]. Additionally, pit latrines with unsealed septic tanks can lead to bacterial contamination of ground water through seepage, which may, in turn, contaminate drinking water sources, particularly when located in close proximity [52]. This is especially problematic in areas with a high-water table such as Bangladesh, where nearly a third of the land area of the country is underwater during rainy seasons.

Little information is available on infection prevention practices during home deliveries. Data from this study revealed gaps in infection prevention practices, particularly related to hand hygiene. Despite the lapses, the limited shared equipment, lack of invasive procedures, and absence of co-dwelling of patients reduce the likelihood of horizontal transmission between households. The more direct route of transmission may be between mothers and their home environment. Although not systematically evaluated in the current study, the majority of the households in this region utilize earthen floors, which allow limited cleaning strategies. These floors permit persistent contamination with human fecal bacteria (such as through tracking by foot traffic or defecation by young children) as well as animal fecal bacteria (who often have a fluid range between indoor and outdoor dwellings and whose feces permeate the household premises). Home births are occurring on these same floors, covered with some bedding materials, providing opportunities for horizontal or vertical transmission of antibiotic-resistant bacteria. While we did not do any environmental sampling of these houses, earthen floors could be an important reservoir for antibiotic-resistant bacteria and should be evaluated in future studies [53,54].

This study has several limitations, with the key limitation being the homogeneity of the population. The similarities in exposures across mothers and newborns limited the ability to detect significant associations, particularly in the setting of ubiquitous colonization. The high colonization prevalence overall demonstrates intense colonization pressure at the community level. Therefore, individual risk factors and colonization outcomes may not be independent, potentially obscuring the identification of meaningful associations as a result of nonlinear disease dynamics.[55] Moreover, the small sample size also limits the capacity to identify risk factors. Additionally, colonization was assigned based on CHROMagar™culture results, which may be an imperfect assessment of colonization. While CHROMagar™ growth may be expected to overcall resistance, reliance on rectal swabs as opposed to stool may reduce the sensitivity [56]. Therefore, true colonization estimates could be higher or lower than reported. However, even with a substantial margin of error, the burden of colonization in this population remains concerningly high. Another limitation is that the data from mothers and birth attendants was self-reported and may be subject to recall or social desirability bias.

## Conclusions

Overall, this study identified alarmingly high rates of ESBL-PB and CRB colonization among mother-newborn pairs undergoing home deliveries in rural Bangladesh, highlighting the critical need for further understanding of community based exposure and transmission. This is especially important in low-resource settings, where socioeconomic factors may play a vital role in the spread of AMR.

## List of abbreviations

AMR: Antimicrobial resistance
GBS: Group B streptococcus
C-section: Cesarean delivery
ESBL-PB: Extended-spectrum beta-lactamase-producing bacteria
CRB: Carbapenem-resistant bacteria
CHAMPS: Child Health and Mortality Prevention Surveillance
WASH: Water, sanitation, and hygiene

## Declarations

### Ethics approval and consent to participate

We obtained written informed consent from all adult study participants. Consent for newborn enrollment was provided by the parents and/or legal guardians. The study protocol was reviewed and approved by the IRB at Stanford University (protocol #53442) and the Research and Ethics Review Committees at icddr,b (protocol #PR-19119).

### Availability of data and materials

All data generated or analyzed during this study are included within the article and its supplementary information files.

## Authors’ contributions

HH contributed to data collection, analysis, and manuscript writing. GSW participated in data analysis and manuscript writing. AS designed the study protocol and contributed to manuscript writing and revisions. MBA, SP, MAI, AP, DZ, ESG, and SL contributed to the conceptual design of the study and interpretation of results and provided substantial edits to the manuscript. MBA and ASS conducted the microbiology methods and contributed to data interpretation. All authors read and approved the final manuscript.

## Acknowledgements

We are grateful to all the participants of this study for providing consent to collect samples and invaluable information. We would also like to thank the CHAMPS Bangladesh team for their collaboration in the use of their surveillance network for identifying births and for all their preceding work to build trust within the community that facilitated participation in the current study. icddr,b acknowledges with gratitude the Governments of Bangladesh and Canada for providing core/unrestricted support.

---

S1 Table 1. Complete list of survey questions and responses of mothers undergoing home-based deliveries, Baliakandi, Bangladesh, 2022 (N=50)

S1 Figure 1. Extended-spectrum beta-lactamase-producing bacterial colonization prevalence by presumptive bacterial identification among mothers and newborns following home-based delivery, Baliakandi, Bangladesh, 2022 (N=50)

S1 Figure 2. Carbapenem-resistant bacterial colonization prevalence by presumptive bacterial identification among mothers and newborns following home-based delivery, Baliakandi, Bangladesh, 2022 (N=50)

S1 Table 2. Birth attendant practices among home-based delivery, Baliakandi, Bangladesh, 2022 (N=9)

